# Strong Genetic Overlaps Between Dimensional and Categorical Models of Bipolar Disorders in a Family Sample

**DOI:** 10.1101/2023.06.24.23291169

**Authors:** Alejandro Arbona-Lampaya, Heejong Sung, Alexander D’Amico, Emma E. M. Knowles, Emily K. Besançon, Ally Freifeld, Ley Lacbawan, Fabiana Lopes, Layla Kassem, Antonio E. Nardi, Francis J. McMahon

## Abstract

**Background:** Bipolar disorder (BD) presents with a wide range of symptoms that vary among relatives, casting doubt on categorical illness models. To address this uncertainly, we investigated the heritability and genetic relationships between categorical and dimensional models of BD in a family sample.

**Methods:** Participants in the Amish-Mennonite Bipolar Genetics (AMBiGen) study were assigned categorical mood disorder diagnoses by structured psychiatric interview and completed the Mood Disorder Questionnaire (MDQ), which assesses lifetime history of manic symptoms and associated impairment. Major MDQ dimensions were analyzed by Principal Component Analysis (PCA) in 726 participants. Heritability and genetic overlaps between categorical diagnoses and MDQ-derived dimensions were estimated with SOLAR-ECLIPSE within 432 genotyped participants.

**Results:** MDQ scores were significantly higher among individuals diagnosed with BD and related disorders, as expected, but varied widely among relatives. PCA suggested a three-component model for the MDQ. Heritability of the MDQ score was 30% (p<0.001), evenly distributed across its three principal components. Strong and significant genetic correlations were found between categorical diagnoses and most MDQ measures.

**Limitations:** Recruitment through probands with BD resulted in increased prevalence of BD in this sample, limiting generalizability. Unavailable genetic data reduced sample size for some analyses.

**Conclusion:** Significant heritability and high genetic correlations between categorical diagnoses and MDQ measures support a genetic continuity between dimensional and categorical models of BD.

## 1. Introduction

Bipolar disorder (BD) is a mental health condition marked by cyclic fluctuations in mood and behavior that impair daily functioning and increase risk for suicide. BD affects up to 4.4% of adults in the United States at some point in their lives (Merikangas et al., 2007). BD can begin with a depressive episode, indistinguishable from unipolar depression, and is frequently unrecognized or misdiagnosed for several years after onset (Carta and Angst, 2016). In addition, many individuals suffer from subclinical symptoms that do not meet categorical diagnostic thresholds (Axelson et al., 2011). Undiagnosed relatives of people with BD report increased mood instability (Stanislaus S et al., 2020). These observations have led to an increasing focus on dimensional measures of psychopathology that may better reflect the full spectrum of illness presentations. (Bruce et al., 2019; Casey et al., 2013; Merikangas et al., 2011; Parker et al., 2016).

The Mood Disorder Questionnaire (MDQ) is a dimensional measure of manic symptoms that is commonly used as a screening tool for BD and related disorders. The MDQ is a self-report questionnaire that assesses 13 manic symptoms, the co-occurrence of these symptoms during distinct episodes, and the degree to which symptoms led to problems with behavior (Hirschfield et al., 2000). Each endorsed symptom constitutes one point for the MDQ Score (MDQS), with scores ranging from zero to 13. The MDQ has been shown to detect probable BD at the widely agreed MDQS of seven or more, with sensitivity around 0.60 and specificity around 0.85 (Wang et al., 2015). Several studies have shown that the MDQ is an effective screening tool for BD in both urban and rural clinical settings (Hardoy et al., 2005; Rouillon et al., 2010; Twiss et al., 2008).

Utilizing the Amish Mennonite Bipolar Genetics Study (AMBiGen) (Gill et al., 2016), we have previously demonstrated that the MDQ is a sensitive and specific screening tool for BD in Anabaptist families ascertained through probands with BD (Dumont et al. 2020). Anabaptists include well-known communities such as the Amish and Mennonites, both of which originated in Western Europe. Anabaptist communities typically comprise large families, well-documented genealogy, rural lifestyles, and a low prevalence of substance abuse (Gill et al., 2016). These characteristics offer advantages in the study of the genetic underpinnings of common disorders like BD.

We have also previously shown that the symptom profiles of manic and depressive episodes are similar between Anabaptist and non-Anabaptist individuals diagnosed with BD (Gill et al., 2016). In the present study, we used the MDQ to investigate the genetic relationship between categorical BD diagnoses and manic symptoms assessed by the MDQ. First, we examined the factor structure of the MDQ by Principal Component Analysis (PCA). These results were compared to published results in other populations. Next, we tested the heritability of the MDQS and scores derived from the PCA to determine which components of the MDQ were most heritable. The analysis gives insight on the extent to which the variance observed in the MDQ components can be attributed to inherited genetic variation. Finally, we tested for genetic correlations between MDQ measures and categorical BD diagnoses in this sample. These analyses reveal to what degree overlapping genetic risk factors contribute to both BD diagnoses and manic symptoms in undiagnosed relatives and could inform future studies aimed at identifying genes that underlie susceptibility to BD in the population.

## 2. Methods

### 2.1. Participants

Participants were recruited as part of the AMBiGen study. In North America, families were recruited through a proband with a diagnosis of BD. We sought to assess all first- or second-degree relatives who were at least 18 years of age. Exclusion criteria included major physical, neurological, or substance use disorders that complicated diagnosis. South American participants all belonged to one of three Mennonite settlements in Brazil who were all descended from a relatively small number of founding couples (Lopes et al., 2016).

Most participants who had an MDQS greater than or equal to seven (Dumont et al. 2020), or who otherwise endorsed a history of mental health problems based on the Past History Schedule (McGuffin et al., 1986) agreed to undergo a direct assessment with the Diagnostic Interview for Genetic Studies (DIGS). The DIGS is a widely-used, semi-structured psychiatric examination that reliably elicits diagnostic criteria for major depression, mania, psychosis, alcohol and drug use, suicidal behavior, and anxiety disorders (Nurnberger et al., 1994). The DIGS assesses lifetime symptoms as well as the most severe periods of major depression and mania. Following the DIGS, two clinicians independently assigned a best estimate final diagnosis (Leckman et al., 1992) based on the interview, available medical records, and reports from relatives.

Categorical diagnoses were grouped into “narrow” and “broad” diagnostic groups based on published family studies (Gershon et al., 1982). Participants diagnosed with bipolar type I (BD1), bipolar type II (BD2) with recurrent depression, and schizoaffective bipolar disorder (SABP) were assigned to the “narrow” group. The “broad” group included the “narrow” group along with BD2 with a single episode of depression, schizoaffective depressive disorder (SAD), recurrent MDD, bipolar disorder not otherwise specified, and schizophrenia (SZ). SZ was included in the “broad” diagnostic group since SZ and BD co-occur in families and share over 80% of common genetic risk factors (reviewed in Gordovez and McMahon, 2020). All other participants, including those with a single episode of major depression (n = 40), were assigned to the “unaffected” diagnostic group. We opted not to incorporate single episodes of major depression and anxiety disorders into the “broad” group since these disorders are very common and do not show strong familial co-aggregation with BD.

A total of 726 participants were included in this study (484 from North America and 242 from Brazil, South America). Of these, 112 were assigned a “narrow” group (BD1 = 89, BD2 recurrent depression = 20, SABP = 3), 212 with a “broad” group (BD1 = 89, BD2 recurrent depression = 20, SABP = 3, BD2 with single episode of depression = 15, MDD (recurrent) = 43, SAD = 2, SZ = 6, other major mood disorders = 34). A total of 514 subjects did not meet the criteria for a “narrow” or “broad” groups, therefore were assigned to the “unaffected” diagnostic group for the purposes of this analysis.

### 2.2. MDQ Screening

As part of the AMBiGen study protocol, most participants were administered the MDQ. This widely used screening tool rates 13 cardinal symptoms of mania, their temporal clustering, and associated impairment. Consistent with the literature, we assigned MDQ scores as a sum of the 13 cardinal symptoms. Thus, scores ranged from 0 to 13. The concurrent symptoms question (CQ) was scored separately: “Of the things we just talked about, have several of these ever happened during the same period of time?” and was converted from Yes or No to 1 or 0. The impairment question (PQ) was also scored separately: “How much of a problem did any of these cause you – like being unable to work; having family, money, or legal troubles; getting into arguments or fights?” and (for purposes of the PCA) was converted from an ordinal scale (“no problem,” “minor problem,” “moderate problem,” and “serious problem”) to 0 for “no problem“ or “minor problem” and 1 for “moderate” or “serious” problem. We also explored the impact of adding the PQ value to the total MDQ score (MDQP), which expanded the maximum score to 14. Prior to statistical analysis, we excluded (n = 50) participants whose MDQ responses contained more than 20% unanswered questions; missing MDQ items among the remaining participants were treated as a zero (no).

### 2.3. Data Analysis

A PCA was conducted using XLSTAT to explore the factor structure of the MDQ in this sample. The Kaiser-Meyer-Olkin test value was 0.93 indicating that this sample is adequate for PCA. Varimax rotations of two and three factors were run to determine which model fit best with the data. Varimax rotation simplifies item loadings by removing the middle ground and clarifying the factor on which data load is based (Dilbeck, 2017). Higher loading values show a factor’s importance in explaining the data, and these values represent the amount of variability in the data that each factor explains.

A genomic relationship matrix (GRM) was created using high-quality single nucleotide polymorphism (SNP) data. The GRM is a key tool in estimating heritability using genomic data, since it enables more accurate estimation of the genetic variance and covariance among individuals, as opposed to relying solely on pedigree information (Jiang et al., 2019).

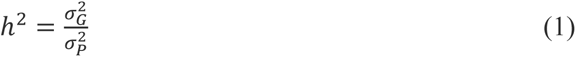

The sample size for the heritability analysis was reduced from 726 to 432 because GRM data was not available for 294 subjects.

Heritability analysis was performed with Solar-Eclipse (v9.0.0) (Kochunov et al., 2019; Seal et al., 2022). Heritability (h^2^) represents the portion of the phenotypic variance (σ^2^) accounted for by additive genetic variance (σ^2^), as seen in **equation 1**. The traits chosen for the heritability analysis were MDQS, CQ, PQ, MDQP, and the rotated components (RC) from the three-factor PCA with varimax rotation (RC1, RC2, and RC3 referring exclusively to the rotated components of the three-factor varimax rotation). Before analysis, all traits underwent an inverse normal transformation to normalize their distributions and facilitate accurate statistical comparisons.

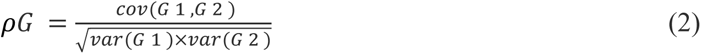

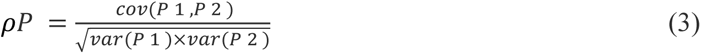

Genetic correlation analyses were performed with SOLAR-Eclipse (Almasy and Blangero, 1998) using the restricted maximum likelihood (REML) method, which estimates the variance-covariance matrix of the genetic effects on pairs of traits. The genetic correlation (ρG) was then calculated as the ratio of the estimated genetic covariance to the square root of the estimated genetic variances for the two traits, providing an estimate of the extent to which the genetic factors underlying the two traits are correlated. This is shown in **equation 2**. The phenotypic correlation (ρP) was estimated with a similar approach, shown in **equation 3**. A ρG value of zero means that the two traits do not share genetic factors, a ρG value of 1 suggests that the genetic factors are entirely shared, and a ρG value of -1 would imply that all the genetic influences on one trait are opposite to those on another trait (Man et al., 2019). ρG values of exactly -1 or 1 should not be overvalued since these values constitute a boundary constraint in the analysis. Results that report a value of -1 or 1 should be considered as close to -1 or 1, but not exactly -1 or 1. Each pairwise correlation of MDQS, CQ, PQ, MDQP, and RC1, RC2, and RC3 with the “narrow” and “broad” groups were examined. To test the significance of the ρG values, we compared the ln likelihood of a restricted null model (with ρG fixed at zero) to that of an alternative model in which the ρG parameter was estimated (Glahn et al., 2012). *P* values <0.05 were considered statistically significant.

## 3. Results

### 3.1. Descriptive Statistics

Sample characteristics are shown in Table 1. MDQS ranged from 0 to 13 (mean = 2.83, SD = 3.49). A total of 109 subjects (15.01%) had an MDQS ≥ 7, the conventional cutoff for screening studies. MDQS scores differed between participants diagnosed with BD1 (mean 5.25, 95% CI ± 0.925) and BD2 (mean 4.46, 95% CI ± 1.24), compared to those with no diagnosis (mean 2.22, 95% CI ± 0.284) (**Figure 1**). In the total sample, the endorsement rate of symptoms on the core 13 items of the MDQ ranged from 7.71% (item 13, “spending money got into trouble”) to 36.36% (item 7, “easily distracted”). As expected, endorsement rates for subjects diagnosed with BD (n = 124) was much higher, ranging from 28.23% (item 13) to 78.23% (item 7). Subjects with a psychiatric diagnosis other than BD (n = 167) had an endorsement rate lower than subjects with a BD diagnosis, but higher than those with no diagnosis (n = 435; details in **Table S1)**.

**Figure 1.**
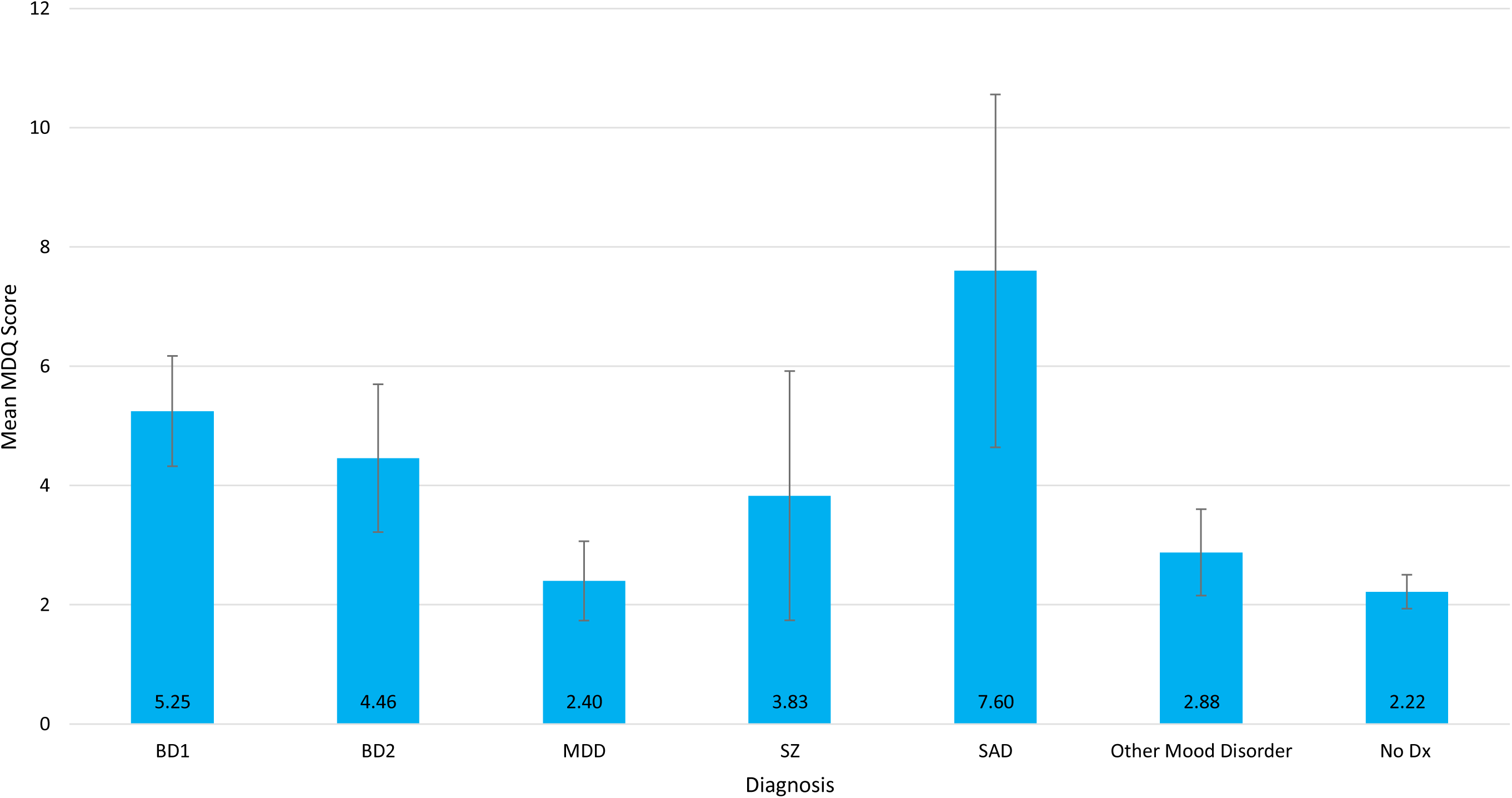
Mean MDQ scores for each mood disorder diagnosis. Bar plots showing the mean Mood Disorder Questionnaire score for each mood disorder diagnosis. Error bars represent 95% CI. Sample sizes can be found in Table 1. BD1 = Bipolar Disorder Type I, BD2 = Bipolar Disorder Type 2, MDD = Major Depressive Disorder, SZ = Schizophrenia, SAD = Schizoaffective Disorder, No dx = No diagnosis.

**Figure 2.**
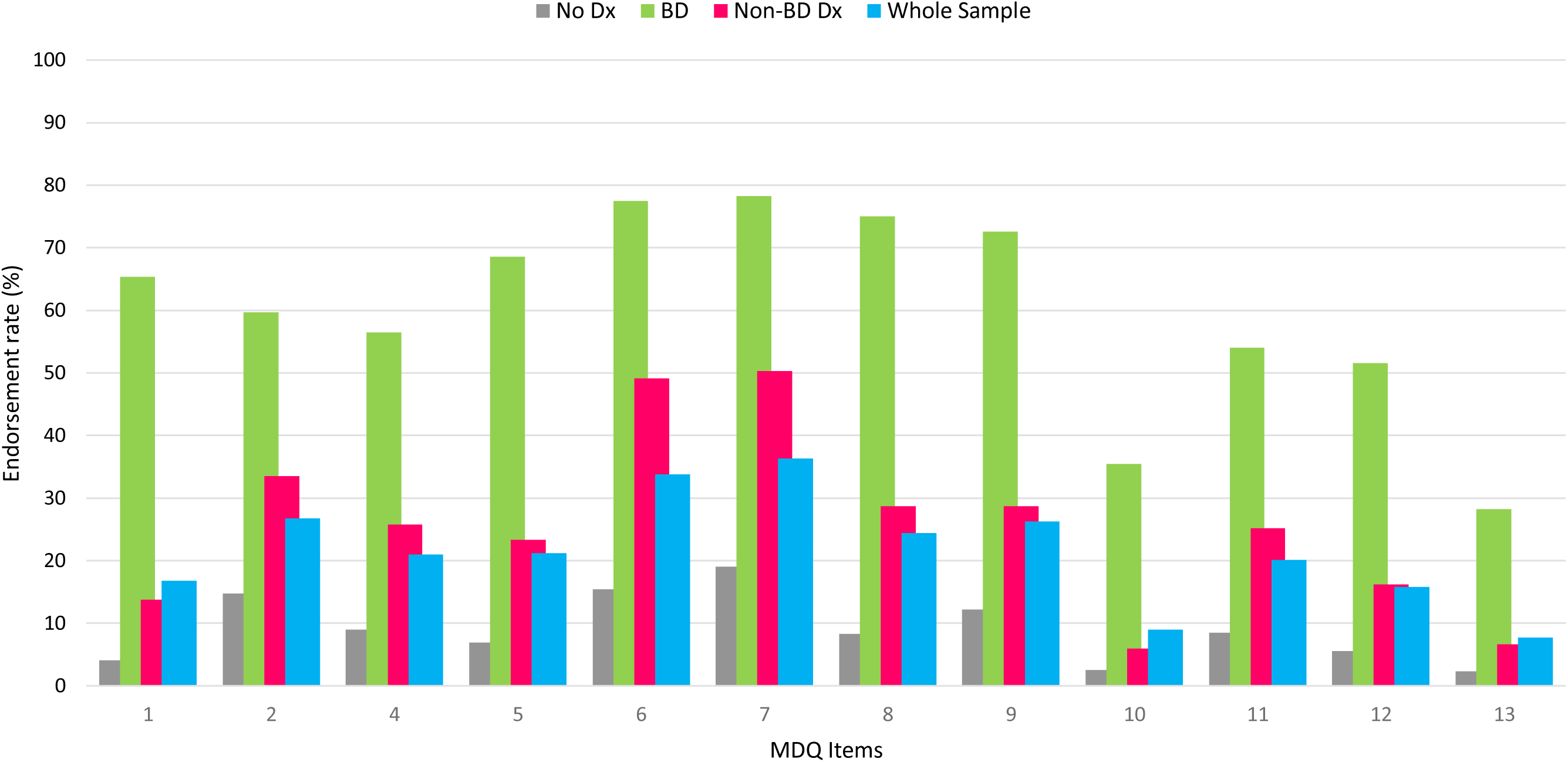
Endorsement rate of the Mood Disorder Questionnaire items. Whole sample (n = 726) is represented by the blue bars, BD sample (n = 124) is represented by the green bars, Non-BD Dx sample (n = 167) is represented by the red bars, No Dx sample (n = 435) is represented by the gray bars. MDQ items: 1 So hyper you get into trouble, 2 Irritable, 3 More self-confident, 4 Less sleep, 5 More talkative, 6 Thoughts raced, 7 Easily distracted, 8 Much more energy, 9 Much more active, 10 Much more social, 11 Much more interested in sex, 12 Excessive, foolish, or risky things, 13 Spending money got into trouble

**Table 1.** Sample characteristics.

| Variables | Total sample (n = 726) |  |
| --- | --- | --- |
| Age in year, mean ± SD, (min, max) | 47 ± 18, | (12-99) |
| Female gender, n (%) | 397 | (54.7) |
| Geographic location, n (%) |  |  |
| North America | 484 | (66.7) |
| Brazil | 242 | (33.3) |
| Diagnosis<sup>†</sup>, n (%) |  |  |
| Bipolar 1 | 89 | (12.3) |
| Bipolar 2 | 35 | (4.8) |
| Major Depressive Disorder | 83 | (11.4) |
| Schizophrenia | 6 | (0.8) |
| Schizoaffective Disorder | 5 | (0.7) |
| Other | 73 | (10.1) |
| No diagnosis | 435 | (59.9) |

Note: SD = Standard Deviation <sup>†</sup> Based on DSM codes

### 3.2. Principal Component Analysis

The PCA revealed that the first two components accounted for 53.05% of the variance in the data. PC1 had an eigenvalue of 5.85 and accounted for 45.05% of the variance while PC2 had an eigenvalue of 1.04 and accounted for 8.00% of the variance. PC3 had an eigenvalue of 0.93, which is below the usual cutoff of 1, but was included in the analysis since it accounted for almost as much variance (7.12%) as PC2 (**Figure S2**). The factor loadings for PC1 were above |0.5| for each item of the MDQ, which indicates a clear and meaningful relationship between the variables in that component. PC2 had two factor loadings above |0.5| for items 2 (irritability) and 7 (distractibility). PC3 had one factor loading above |0.5|, for overspending (**Table S3**).

Since PC1 was not highly differentiated in the factor loadings, we explored varimax rotation with two and three PCs. The PCA with a two-factor varimax rotation maintained the 53.05% variance among the two RCs, but this was spread more evenly across the first two PCs (34.01% and 19.04%, respectively) and factor loadings were more differentiated (**Table S4**). The PCA with a three-factor varimax rotation captured 60.18% of the variance among RC, RC2 and RC3 (**Figure 3**). Factor loadings were well balanced among all 3 RCs (**Figure 4**). Results of the two-factor varimax rotation were similar (Table S4)

**Figure 3.**
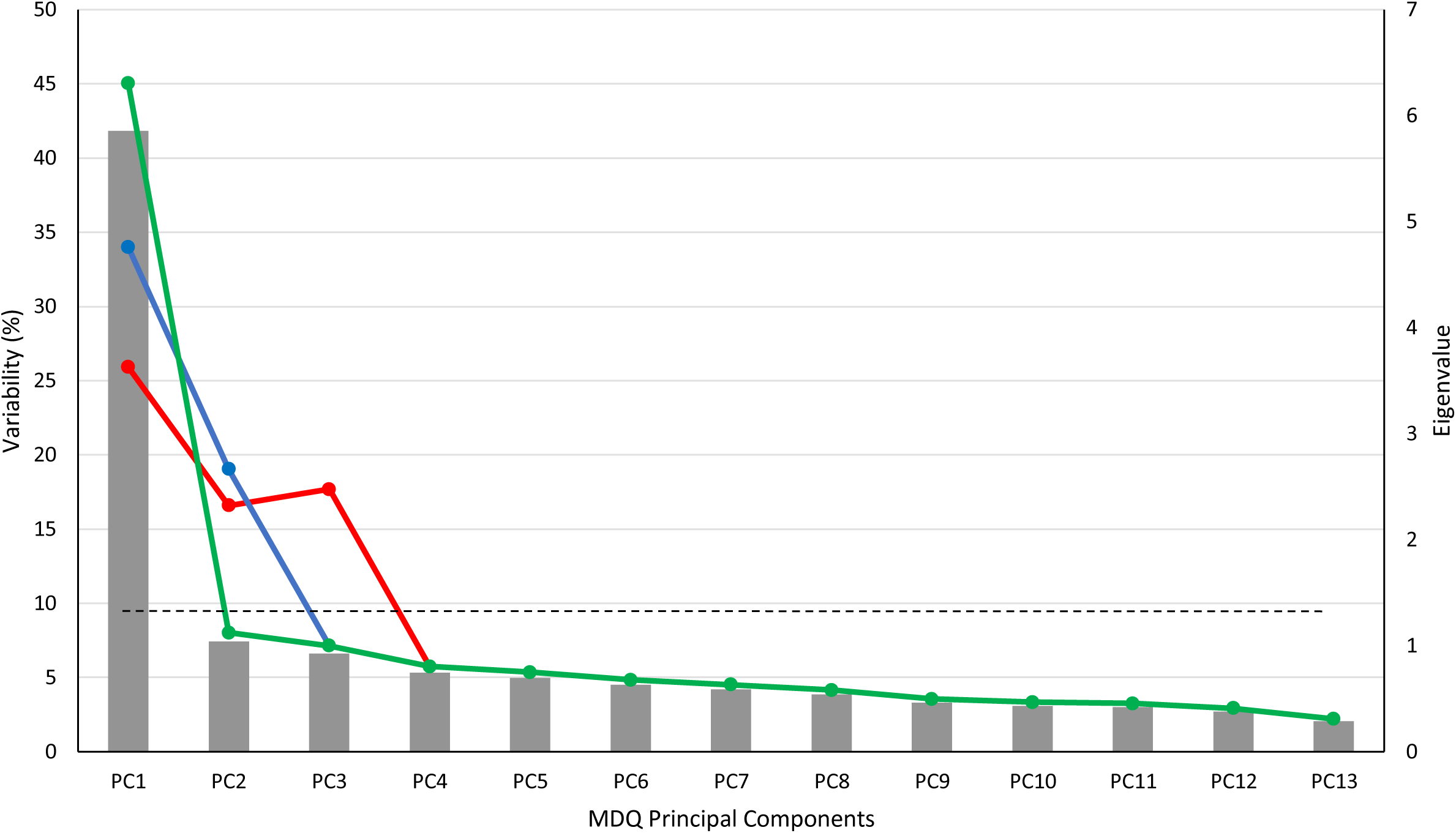
Scree Plot comparing the variability percentage between Principal Component Analyses. Eigenvalues are shown a gray bar. The green line represents the variability (%) of the PCA with no varimax rotation. The blue line represents the variability (%) of the PCA with a two-factor varimax rotation The red line represents the variability (%) of the three-factor varimax rotation PCA variability. The dotted line represents where eigenvalues = 1.0. PC = principal component

**Figure 4.**
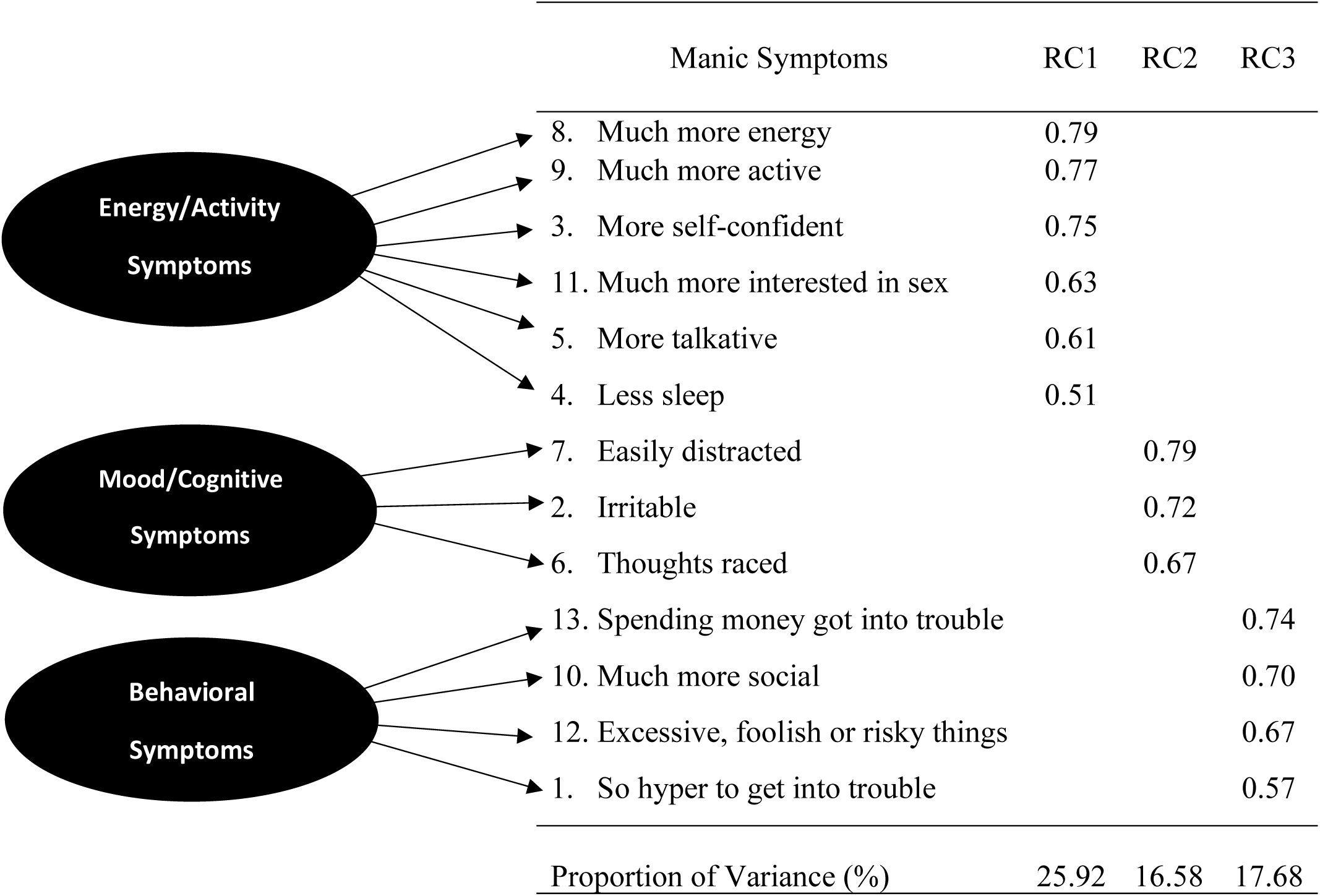
Three-factor varimax rotation Principal Component Analysis model with factor loading scores for each rotated component. Factor loadings > 0.5 are shown. RC = rotated component.

### 3.3. Heritability analysis

Results of the heritability analyses are shown in **Table 2**. The MDQP and the MDQS had the highest h^2^, 36% and 30% respectively, followed by the PQ with 26%; all were significantly different from zero (p ≤ 0.001). The h^2^ for PC3 was 18% (p ≤ 0.05) (**Table S6**). PC1 and PC2 did not show statistically significant h^2^ values despite representing the most variance. These results prompted further analysis via the varimax rotations. Similar to the analysis of the principal components, the h^2^ of the first RC of the two-factor varimax rotation did not differ statistically from zero, while the second RC did (**Table S6**). In contrast, RC1 and RC2 both showed statistically significant h^2^.

**Table 2.** Heritability Estimates and Correlation Analysis for Dimensional and Categorical Measures.

| Traits | $h^2$ (Std. E) | $\rho_G$ (Std E.) | $\rho_P$ | p-value | |
| --- | --- | --- | --- | --- | --- |
| MDQS | 0.30 (0.10) | - | - | <0.001 |  |
| CQ | 0.09 (0.08) | - | - | >0.05 |  |
| PQ | 0.27 (0.10) | - | - | <0.001 |  |
| MDQP | 0.36 (0.11) | - | - | <0.001 |  |
| RC1 | 0.21 (0.10) | - | - | <0.05 |  |
| RC2 | 0.20 (0.09) | - | - | <0.05 |  |
| RC3 | 0.13 (0.13) | - | - | >0.05 |  |
| Correlations | | | | $\rho_G$ p-value | $\rho_P$ p-value |
| MDQS-N | - | 1.00<sup>†</sup> | 0.16 | >0.05 | <0.01 |
| MDQS-B | - | 0.90 (0.22) | 0.21 | <0.01 | <0.001 |
| PQ-N | - | 1.00<sup>†</sup> | 0.37 | <0.01 | <0.001 |
| PQ-B | - | 0.64 (0.25) | 0.40 | <0.05 | <0.001 |
| MDQP-N | - | 1.00<sup>†</sup> | 0.23 | >0.05 | <0.001 |
| MDQP-B | - | 0.95 (0.10) | 0.28 | <0.001 | <0.001 |
| RC1-N | - | 1.00<sup>†</sup> | 0.32 | >0.05 | <0.001 |
| RC1-B | - | 0.62 (0.31) | 0.37 | >0.05 | <0.001 |
| RC2-N | - | 1.00<sup>†</sup> | 0.30 | >0.05 | <0.001 |
| RC2-B | - | 1.00<sup>†</sup> | 0.42 | >0.05 | <0.001 |
| RC3-N | - | 1.00<sup>†</sup> | 0.35 | >0.05 | <0.001 |
| RC3-B | - | 1.00<sup>†</sup> | 0.11 | <0.05 | <0.001 |

Note: MDQS = MDQ score, CQ = concurrent symptoms question (“...have several of these ever happened during the same period of time?”) PQ = impairment question (“How much of a problem did any of these cause you...?”), MDQP = MDQS + PQ, RC1 = first rotated component, RC2 = second rotated component, RC3 = third rotated component, N = “narrow” categorical diagnosis group, B = “broad” categorical diagnosis group. $\rho_G$ = genotypic correlation, $\rho_P$ = phenotypic correlation. <sup>†</sup> Standard error not computable. Note that $\rho_G$ values of 1.00 represent a boundary constraint in the analysis method and should not be construed as complete genetic overlap between traits.

### 3.4. Genetic and Phenotypic Correlations

Genetic overlaps between MDQ traits and categorical diagnoses were estimated as genetic correlations (**Table 3)**. While all genetic correlations were above 0.6, values for the “narrow” diagnosis group were significantly different from 0 only for impairment (PQ). The “broad” diagnostic group showed strong and significant genetic correlations with most MDQ-derived traits Phenotypic correlations, while smaller than genetic correlations, were all significantly different from zero.

## 4. Discussion

This is the first study to investigate heritability of the MDQ in families and genetic overlap with categorical BD diagnoses. Studying the heritability of the MDQ can help us better understand the genetic underpinnings of BD and related mood disorders. By identifying the extent to which the MDQ is influenced by genetic factors, we can gain insight into heritable mechanisms that contribute to manic symptoms.

The MDQS was shown to be significantly heritable (30%) (**Table 2**), although lower than most estimates of the heritability of categorical BD diagnosis (40-85%) (Bruce et al., 2019; Fabbri, 2020; Lee et al., 2013; McGuffin et al., 2003). The PQ was also significantly heritable (26%) (**Table 2**). The MDQP showed the highest heritability of the dimensional traits we tested (36%) (**Table 2**). Given that the MDQP is an exploratory trait, this result should be viewed with caution. The higher heritability of the MDQP may reflect the increased specificity and reduced noise associated with this phenotype, as well as the extra information it conveys. Further studies are needed to validate MDQP as a dimensional measure of “bipolarity”, however.

Previous MDQ studies have shown that either a two factor (Carta et al., 2014; Chung et al., 2008; Ouali et al., 2020; Sanchez-Moreno et al., 2008) or three factor (Chung et al., 2009; Jon et al., 2009; Massidda et al., 2016; Mundy et al., 2023; Yang et al., 2011) best fit the data structure of the MDQ. We put this to the test by performing a PCA with varimax rotation on both a two-factor and a three-factor model, comparing the results to a PCA without varimax rotation. The three-factor model performed slightly better in this sample. These findings align with similar three-factor models observed in Korean (Jon et al., 2009), Chinese (Yang et al., 2011), Hong Kong (Chung et al., 2009), and UK (Mundy et al., 2023) populations, suggesting a cross-cultural consistency in the underlying dimensions of the MDQ. While specific dimension labels may differ slightly, the overall three-factor model (**Figure 3**) seems robust, consistent with core dimensions that capture typical constructs of mania.

The heritability estimates for PC1, PC2, and PC3 were found to be modest; only PC2 and PC3 demonstrated statistical significance (**Table S6**). This likely reflects reduced power due to sample size. The larger sample size available for the PCA – which did not rely only on genotyped individuals - provided more statistical power. These findings are in line with expectations given that PC2 and PC3 are subcomponents of the MDQ, and their joint contribution approximates, but does not fully capture the heritability of the MDQ. We also examined the heritability of the two-factor and three-factor RC. In the two-factor model, only the second RC showed statistically significant heritability. In contrast, RC1 and RC2 showed significant heritability, while RC3 did not. These results suggest that energy, self-confidence, and expansive mood, which load on RC1 and RC2 are more heritable than the behavioral symptoms that load on RC3. However, behavioral symptoms of mania were endorsed by relatively few participants in this sample (**Figure 2)**, which probably reflects cultural and environmental factors characteristic of the Anabaptist community we studied.

Our study identified a significant genetic and phenotypic correlations between MDQS and the “broad” diagnostic group in this sample (ρG = 0.90, ρP = 0.21). A genetic correlation of this magnitude suggests that the two traits share most genetic risk factors, while the lower value for the phenotypic correlation suggests that these genetic risks are expressed differently in different individuals. A similar relationship was found between MDQP and the “broad” diagnostic group (ρG = 0.95 and ρP = 0.28) and between PQ and the “narrow” group. These genetic correlations are higher than those reported between SZ and BD (ρG = 0.68) and between BD and MDD (ρG = 0.47) (Fabbri, 2021).

To our knowledge only two previous studies have examined the relationship between dimensional and categorical models of BD. Bruce et al. (2019) found that a quantitative measure of bipolar symptoms was significantly heritable and concluded that “bipolarity trait assessment may be used to supplement the diagnosis of BD in future genetic studies and could be especially useful for capturing subclinical genetic contributions to a BD phenotype.” Those results largely agree with the present study, even though we employed a different measure of “bipolarity.” Recently, Mundy et al. (2023) conducted a genome-wide association study of MDQ scores on a sample of the UK population. The purpose of their study was to determine the validity of the MDQ as a screening tool for BD in at-risk populations. The authors found that the MDQ was not significantly heritable and was most strongly associated with symptoms of general distress or psychopathology, not just manic symptoms, in that population sample. These results contrast with the significant heritability of the MDQS and genetic overlap with BD in the present study. This likely reflects differences in the study population. Our study was based on individuals ascertained through family members with BD. Manic symptoms reported by relatives of people with BD are more likely to share common genetic determinants with BD than manic symptoms reported by the general population.

## 5. Limitations

This study has several limitations. First, because of unavailable genetic data, the heritability and genetic correlation analyses had a reduced sample size, which decreased power to detect weak effects and may affect precision and generalizability of the apparently stronger effects. Second, the recruitment of participants through probands with BD resulted in a higher prevalence of BD in this sample, reducing the generalizability of the genetic associations we report to populations with lower baseline rates of BD. Third, while this study sheds light on the genetic relationships between dimensional and categorical measures of BD, it does not point to specific genes. Nevertheless, the results support the value of the MDQ as a screening tool, especially among individuals at high a-priori risk of BD.

## Data Availability

All data produced in the present study are available upon reasonable request to the authors.

## Author Contributions

AAL and FJM designed the study. EKB, AEN, FL, AD, and AF acquired the data. EKB, LK, LL, and FJM conducted patient interviews. AAL, HS, and FJM analyzed the data. AAL drafted the manuscript. All the authors contributed to the editing of the manuscript and have approved the final version.

## Acknowledgements

All participants provided informed consent under protocol 80-M-0082. We thank the participants and their families for contributing to this study. This work utilized the computational resources of the NIH HPC Biowulf cluster (http://hpc.nih.gov).

