## Supplementary Tables for "Strong Genetic Overlaps Between Dimensional and Categorical Models of Bipolar Disorders in a Family Sample"

**Table S1.** Descriptive statistics for the MDQ items

|  | Whole Sample  (n = 726) | | BD  (n = 124) | | Non-BD Dx  (n = 167) | | No Dx  (n = 435) | |
| --- | --- | --- | --- | --- | --- | --- | --- | --- |
| MDQ Items | **Yes** | **Endorsement rate (%)** | **Yes** | **Endorsement rate (%)** | **Yes** | **Endorsement rate (%)** | **Yes** | **Endorsement rate (%)** |
| 1 | 122 | 16.80 | 81 | 65.32 | 23 | 13.77 | 18 | 4.13 |
| 2 | 194 | 26.72 | 74 | 59.68 | 56 | 33.53 | 64 | 14.71 |
| 3 | 177 | 24.38 | 87 | 70.16 | 43 | 25.75 | 47 | 10.80 |
| 4 | 152 | 20.94 | 70 | 56.45 | 43 | 25.75 | 39 | 8.97 |
| 5 | 154 | 21.21 | 85 | 68.55 | 39 | 23.35 | 30 | 6.90 |
| 6 | 245 | 33.75 | 96 | 77.42 | 82 | 49.10 | 67 | 15.40 |
| 7 | 264 | 36.36 | 97 | 78.23 | 84 | 50.30 | 83 | 19.08 |
| 8 | 177 | 24.38 | 93 | 75.00 | 48 | 28.74 | 36 | 8.27 |
| 9 | 191 | 26.31 | 90 | 72.58 | 48 | 28.74 | 53 | 12.18 |
| 10 | 65 | 8.95 | 44 | 35.48 | 10 | 5.99 | 11 | 2.53 |
| 11 | 146 | 20.11 | 67 | 54.03 | 42 | 25.15 | 37 | 8.50 |
| 12 | 115 | 15.84 | 64 | 51.61 | 27 | 16.17 | 24 | 5.52 |
| 13 | 56 | 7.71 | 35 | 28.23 | 11 | 6.59 | 10 | 2.23 |

Note: Dx = diagnosis, Whole sample (n = 726), BD (n = 124), Non-BD Dx (n = 167), No Dx (n = 435). MDQ items 1 So hyper you get into trouble, 2 Irritable, 3 More self-confident, 4 Less sleep, 5 More talkative, 6 Thoughts raced, 7 Easily distracted, 8 Much more energy, 9 Much more active, 10 Much more social, 11 Much more interested in sex, 12 Excessive, foolish, or risky things, 13 Spending money got into trouble

Table **S2.** Pairwise Pearson Correlation matrix of the MDQ items

|  | Q13 | Q4 | Q10 | Q12 | Q11 | Q1 | Q5 | Q3 | Q8 | Q9 | Q2 | Q6 | Q7 |
| --- | --- | --- | --- | --- | --- | --- | --- | --- | --- | --- | --- | --- | --- |
| Q13 | 1.00 |  |  |  |  |  |  |  |  |  |  |  |  |
| Q4 | 0.30 | 1.00 |  |  |  |  |  |  |  |  |  |  |  |
| Q10 | 0.34 | 0.38 | 1.00 |  |  |  |  |  |  |  |  |  |  |
| Q12 | 0.38 | 0.33 | 0.48 | 1.00 |  |  |  |  |  |  |  |  |  |
| Q11 | 0.32 | 0.37 | 0.41 | 0.42 | 1.00 |  |  |  |  |  |  |  |  |
| Q1 | 0.39 | 0.39 | **0.52** | **0.55** | 0.41 | 1.00 |  |  |  |  |  |  |  |
| Q5 | 0.32 | 0.45 | 0.44 | 0.48 | 0.46 | **0.57** | 1.00 |  |  |  |  |  |  |
| Q3 | 0.26 | 0.36 | 0.37 | 0.34 | 0.46 | 0.47 | **0.51** | 1.00 |  |  |  |  |  |
| Q8 | 0.33 | 0.48 | 0.43 | 0.46 | **0.52** | **0.55** | **0.61** | **0.58** | 1.00 |  |  |  |  |
| Q9 | 0.32 | 0.40 | 0.38 | 0.40 | **0.53** | 0.46 | **0.51** | 0.49 | **0.68** | 1.00 |  |  |  |
| Q2 | 0.23 | 0.22 | 0.20 | 0.33 | 0.29 | 0.35 | 0.38 | 0.27 | 0.34 | 0.31 | 1.00 |  |  |
| Q6 | 0.28 | 0.41 | 0.30 | 0.41 | 0.36 | 0.46 | 0.44 | 0.35 | 0.45 | 0.38 | 0.34 | 1.00 |  |
| Q7 | 0.25 | 0.31 | 0.24 | 0.36 | 0.37 | 0.42 | 0.41 | 0.34 | 0.38 | 0.32 | 0.41 | **0.53** | 1.00 |

Note: All values were different from 0 with a significance level alpha=0.05. MDQ items: Q1 So hyper you get into trouble, Q2 Irritable, Q3 More self-confident, Q4 Less sleep, Q5 More talkative, Q6 Thoughts raced, Q7 Easily distracted, Q8 Much more energy, Q9 Much more active, Q10 Much more social, Q11 Much more interested in sex, Q12 Excessive, foolish, or risky things, Q13 Spending money got into trouble.

**S3.** Heat map of the factor loadings of the Principal Component Analysis of the Mood Disorder Questionnaire items. (n = 726)

| Manic Symptoms | PC1 | PC2 | PC3 |
| --- | --- | --- | --- |
| 1. So hyper to get into trouble | 0.76 | -0.01 | 0.21 |
| 2. Irritable | 0.52 | 0.54 | -0.02 |
| 3. More self-confident | 0.68 | -0.20 | -0.34 |
| 4. Less sleep | 0.62 | -0.11 | -0.05 |
| 5. More talkative | 0.77 | -0.03 | -0.10 |
| 6. Thoughts raced | 0.65 | 0.40 | -0.02 |
| 7. Easily distracted | 0.61 | 0.57 | -0.05 |
| 8. Much more energy | 0.80 | -0.18 | -0.27 |
| 9. Much more active | 0.72 | -0.22 | -0.30 |
| 10. Much more social | 0.63 | -0.33 | 0.35 |
| 11. Much more interested in sex | 0.69 | -0.15 | -0.15 |
| 12. Excessive, foolish or risky things | 0.69 | -0.00 | 0.38 |
| 13. Spending money got into trouble | 0.53 | -0.08 | 0.54 |
| Proportion of Variance (%) | 45.05 | 8.00 | 7.12 |

Note: PC = principal component

**S4.** Heat map of the factor loadings of the two-factor varimax rotation Principal Component Analysis of the Mood Disorder Questionnaire items. (n = 726)

| Manic Symptoms | RC1 | RC2 |
| --- | --- | --- |
| 1. So hyper to get into trouble | 0.65 | 0.41 |
| 2. Irritable | 0.14 | 0.73 |
| 3. More self-confident | 0.67 | 0.21 |
| 4. Less sleep | 0.58 | 0.25 |
| 5. More talkative | 0.66 | 0.39 |
| 6. Thoughts raced | 0.33 | 0.69 |
| 7. Easily distracted | 0.20 | 0.81 |
| 8. Much more energy | 0.77 | 0.29 |
| 9. Much more active | 0.73 | 0.21 |
| 10. Much more social | 0.71 | 0.07 |
| 11. Much more interested in sex | 0.65 | 0.25 |
| 12. Excessive, foolish or risky things | 0.58 | 0.37 |
| 13. Spending money got into trouble | 0.48 | 0.23 |
| Proportion of Variance (%) | 34.01 | 19.04 |

Note: PC = principal component

**S5.** Heat map of the factor loadings of the three-factor varimax rotation Principal Component Analysis of the Mood Disorder Questionnaire items. (n = 726)

| Manic Symptoms | RC1 | RC2 | RC3 |
| --- | --- | --- | --- |
| 1. So hyper to get into trouble | 0.42 | 0.35 | 0.57 |
| 2. Irritable | 0.16 | 0.72 | 0.12 |
| 3. More self-confident | 0.75 | 0.17 | 0.13 |
| 4. Less sleep | 0.51 | 0.21 | 0.31 |
| 5. More talkative | 0.61 | 0.35 | 0.33 |
| 6. Thoughts raced | 0.30 | 0.67 | 0.23 |
| 7. Easily distracted | 0.22 | 0.79 | 0.14 |
| 8. Much more energy | 0.79 | 0.24 | 0.24 |
| 9. Much more active | 0.77 | 0.17 | 0.20 |
| 10. Much more social | 0.38 | -0.00 | 0.70 |
| 11. Much more interested in sex | 0.63 | 0.21 | 0.28 |
| 12. Excessive, foolish or risky things | 0.26 | 0.31 | 0.67 |
| 13. Spending money got into trouble | 0.09 | 0.17 | 0.74 |
| Proportion of Variance (%) | 25.92 | 16.58 | 17.68 |

Note: PC = principal component

**S6.** Heritability estimates for MDQS, CQ, PQ, MDQP, Narrow, Broad, PC1, PC2, and PC3, 2F-RC1, 2F-RC2, 3F-RC1, 3F-RC2, and 3F-RC3.

| Traits | n | h^2^ | Std. E | p-value |
| --- | --- | --- | --- | --- |
| MDQS | 432 | 0.30 | 0.10 | <0.001 |
| CQ | 432 | 0.09 | 0.08 | 0.12 |
| PQ | 432 | 0.27 | 0.10 | <0.001 |
| MDQP | 432 | 0.36 | 0.11 | <0.001 |
| Narrow | 432 | 0.08 | 0.19 | 0.34 |
| Broad | 432 | 0.28 | 0.14 | <0.05 |
| PC1 | 432 | 0.12 | 0.09 | 0.057 |
| PC2 | 432 | 0.13 | 0.09 | 0.054 |
| PC3 | 432 | 0.18 | 0.10 | <0.05 |
| 2F-RC1 | 432 | 0.02 | 0.07 | 0.37 |
| 2F-RC2 | 432 | 0.21 | 0.09 | <0.01 |
| 3F-RC1 | 432 | 0.15 | 0.10 | <0.05 |
| 3F-RC2 | 432 | 0.18 | 0.09 | <0.05 |
| 3F-RC3 | 432 | 0.13 | 0.09 | 0.056 |

Note: MDQS = Mood Disorder Questionnaire score, CQ = concurrent symptoms question (“…have several of these ever happened during the same period of time?”) PQ = impairment question (“How much of a problem did any of these cause you…?”), MDQP = MDQS + PQ, PC = principle component, 2F-RC = two-factor varimax rotation component, 3F-RC = three-factor varimax rotation component.

**S7.** Correlation estimates of the phenotypes with the categorical diagnoses

| Traits | ρG | ρP |
| --- | --- | --- |
| MDQS-N | 1.00^†^ | 0.16^**^ |
| MDQS-B | 0.90^**^ | 0.21^***^ |
| MDQS-PQ | 0.41 | 0.02 |
| CQ-N | 1.00^†^ | 0.47^***^ |
| CQ-B | 0.84 | 0.58^***^ |
| PQ-N | 1.00^**†^ | 0.37^***^ |
| PQ-B | 0.64^*^ | 0.40^***^ |
| MDQP-N | 1.00^†^ | 0.23^***^ |
| MDQP-B | 0.95^***^ | 0.28^***^ |
| RC1-N | 1.00^†^ | 0.32^***^ |
| RC1-B | 0.62 | 0.37^***^ |
| RC2-N | 1.00^†^ | 0.30^***^ |
| RC2-B | 1.00^†^ | 0.42^***^ |
| RC3-N | 1.00^†^ | 0.35^***^ |
| RC3-B | 1.00^**†^ | 0.11^***^ |

Note: ρG = genotypic correlation, ρP = phenotypic correlation. MDQS = MDQ score, CQ = concurrent symptoms question (“…have several of these ever happened during the same period of time?”) PQ = impairment question (“How much of a problem did any of these cause you…?”), MDQP = MDQS + PQ, RC1 = first rotated component, RC2 = second rotated component, RC3 = third rotated component * p ≤ 0.05 ** p ≤ 0.01 *** p ≤ 0.001. P-values indicate correlations that are significantly different from zero. † Standard error not computable.
