## Supplementary Figures for "Strong Genetic Overlaps Between Dimensional and Categorical Models of Bipolar Disorders in a Family Sample"

| Number of MDQ symptoms endorsed | Number of subjects |
| --- | --- |
| 0 | 280 |
| 1 | 78 |
| 2 | 75 |
| 3 | 71 |
| 4 | 51 |
| 5 | 28 |
| 6 | 26 |
| 7 | 19 |
| 8 | 21 |
| 9 | 18 |
| 10 | 27 |
| 11 | 8 |
| 12 | 11 |
| 13 | 15 |

**Figure S1.** Bar plot showing the distribution of Mood Disorder Questionnaire (MDQ) scores for the whole sample (n = 726)

Eigenvalue

Cumulative variability (%)

| MDQ Principal Components | Eigenvalue |
| --- | --- |
| PC1 | 5.85671 |
| PC2 | 1.04049 |
| PC3 | 0.926164 |
| PC4 | 0.746137 |
| PC5 | 0.695629 |
| PC6 | 0.629243 |
| PC7 | 0.586726 |
| PC8 | 0.539119 |
| PC9 | 0.460476 |
| PC10 | 0.433022 |
| PC11 | 0.420871 |
| PC12 | 0.379083 |
| PC13 | 0.28633 |

|  | Cumulative % |
| --- | --- |
| PC1 | 45.0516 |
| PC2 | 53.0554 |
| PC3 | 60.1797 |
| PC4 | 65.9192 |
| PC5 | 71.2702 |
| PC6 | 76.1106 |
| PC7 | 80.6238 |
| PC8 | 84.7709 |
| PC9 | 88.313 |
| PC10 | 91.644 |
| PC11 | 94.8814 |
| PC12 | 97.7975 |
| PC13 | 100 |

**Figure S2**. Plot of the eigenvalues based on the data from the Principal Component Analysis. Eigenvalues are shown as bar plots in blue. Cumulative variability % is shown as a red line. PC = principal component
